# A Generalized Evolutionary Classifier for Evolutionary Guided Precision Medicine

**DOI:** 10.1101/2020.09.24.20201111

**Authors:** Matthew McCoy, Chen-Hsiang Yeang, Deepak Parashar, Robert A. Beckman

## Abstract

**Background:** Current precision medicine (CPM) matches patients to therapies, utilizing traditional biomarker classifiers. Dynamic precision medicine (DPM) is an evolutionary guided precision medicine (EGPM) approach that specifically accounts for intratumoral genetic heterogeneity and evolutionary dynamics. DPM adapts as frequently as every six weeks, plans proactively for future resistance development, and incorporates multiple therapeutic agents. Simulations indicate that DPM can double mean and median survival and significantly improve long-term survival in a cohort of 3 million virtual patients representing a pan-oncology spectrum. Given the cost and invasiveness of monitoring subclones frequently in the DPM paradigm, we sought to determine the value of a DPM window study of only two six-week adaptations (“moves”).

**Methods:** 3 million virtual patients, differing in DPM input parameters of initial subclonal compositions, drug sensitivities, and growth and mutational kinetics, were simulated as previously described. Each virtual patient was treated with CPM, DPM, and DPM for two moves followed by CPM.

**Results:** The first two DPM moves provide similar benefit to a five-year, 40-move sequence in the full virtual population. In simulations, if the first two moves are identical for DPM and CPM, patients will not benefit from DPM (89% negative predictive value). The patient subset (20%) in which two-move and 40-move DPM sequences are closely similar in outcome has extraordinary predicted benefit (HR-DPM/CPM 0.04).

**Conclusions:** The first two DPM moves provide most of the clinical benefit of DPM, reducing the duration of required subclonal monitoring. This leads to an evolutionary classifier (EC) that selects patients who will benefit, i.e. those in whom DPM and CPM recommendations differ early. This EC development paradigm may apply to other EGPM approaches despite different underlying assumptions.

**CONTEXT SUMMARY:**

- Key objective: Current precision medicine (CPM) performs static matching of biomarker classifiers to therapies. We asked, using computer simulation, whether dynamic precision medicine (DPM), a highly adaptive and proactive evolutionary guided precision medicine (EGPM) paradigm, could benefit patients with only two six-week adaptive treatment periods (“moves”), and still enhance long term survival by preventing late term relapses.
- Knowledge generated: Two moves of DPM were highly effective, nearly as effective as 40 moves in the full population. Patients for whom DPM and CPM recommend the same treatment sequence for the first two moves will likely not benefit from DPM.
- Relevance: A 2-move DPM paradigm is far more cost-effective and less invasive than a 40-move paradigm, and opens up the neoadjuvant period for studying DPM. The findings establish an evolutionary classifier (EC) for selecting patients who will benefit from DPM. This general approach to developing an EC may work for other EGPM.

## INTRODUCTION

Current precision cancer medicine matches therapies to static, consensus molecular patterns. Patients are treated with a therapy for as long as they are benefiting, and upon progression and relapse the process is repeated with a different therapy. This approach has resulted in substantial patient benefits and is a major current direction in oncology. However, despite the benefits of this approach, duration of response is variable, and long-term remissions and cures remain elusive.

Cancers are constantly evolving subclonal heterogeneity. Ultradeep sequencing (20,000X) of colorectal cancers at diagnosis and a novel approach to modeling the evolution of very rare subclones indicate that any cancer containing enough cells to be detectable will have at least one cell with a resistance mutation to any single therapy, and that as the cancer burden increases during a clinical course, cells may evolve that are simultaneously resistant to multiple elements of a therapeutic cocktail, at a rate substantially faster than previously anticipated^1, 2^. Rare resistant cells may grow out relatively free of competition from other cancer cells in numerous micrometastases that are small enough to allow ready diffusion of oxygen and nutrients. Infiltration of major organs with metastatic disease, rather than growth of large primary lesions, generally leads to cancer mortality^1, 3^.

Explicit consideration of subclonal heterogeneity and evolutionary dynamics may improve clinical results by preventing the evolution of resistance. Several strategies have been proposed for incorporating evolutionary dynamics in a personalized manner^4-7^. We term these ***evolutionary guided precision medicine (EGPM)*** strategies. A strategy is defined not as a particular therapy or regimen, but rather as an algorithm for determining optimal therapy sequences on an individual basis.

Our work has been focused on ***dynamic precision medicine (DPM)***, a specialized EGPM that explicitly considers subclonal heterogeneity and dynamics, adapts very frequently, and proactively plans ahead based on estimated risks of future events^6^. DPM, unlike earlier EGPMs, considers optimal sequencing of multiple non-cross resistant therapies, rather than focusing on optimal scheduling of a single therapy. Extensive simulation has shown significant improvements in relapse prevention and a doubling of median survival^6^. Primary resistance may be due to non-genetic plasticity including gene regulation or to pre-existing mutations, whereas later relapses will increasingly be due to epigenetic and genetic sub-clonal evolution, the latter including not only mutations but also copy number changes and other rearrangements.

DPM explicitly considers minority subclones and heritable genetic and epigenetic evolutionary dynamics, with the goal of maximizing survival by balancing treating current disease and preventing refractory disease relapse, the latter accomplished by eliminating small singly-resistant subclones before they can evolve sub-subclonal variants simultaneously resistant to multiple non-cross resistant agents. DPM predicts that “hypermutator subclones” with a higher mutation rate will arise due to random mutations in proteins responsible for genome integrity, such as replication and repair proteins^6, 8^. These hypermutator subclones can rapidly evolve cells that are simultaneously resistant to all the agents in a therapeutic cocktail, and are a particular priority for early elimination. The DPM prediction that hypermutator subclones will be enriched in resistant samples is being verified using a fluorescent reporter assay for hypermutators as well as ultradeep DNA sequencing^9^. As a result, DPM will often recommend brief periods of prioritizing elimination of rare hypermutator subclones vs. debulking the tumor.

DPM changes therapy very frequently and plans ahead for potential future evolution of rare hypermutator subclones and others using an adaptive evolutionary model to predict the optimal treatment regimen within a short time window - e.g. two 3-week therapy cycles, although the length of this window can be adjusted. We term each therapy adaptation a “move” in analogy to chess. Frequent changes in therapy are a hallmark of DPM because this minimizes the constant, predictable selection pressure from treatment with a constant single or combination therapy until progression or relapse.

Instead, frequent adaptation complicates evolutionary pathways to multiple resistance^10^. DPM considers the probabilities of distant outcomes up to 5 years in the future when determining each move, and may be superior at prioritizing these longer timescales of interest vs. other algorithms that focus on short term outcomes such as tumor shrinkage^11^.

In an extensive simulation of 3 million virtual patients, where each patient represented a unique starting state of prevalence of sensitive and resistant subclones, and a unique set of parameters including net growth rates, phenotypic transition rates of each subclone between sensitivity and resistance to two non-cross resistant therapies, and sensitivity and resistance values, DPM doubled median survival compared to ***current precision medicine (CPM;*** *targeted therapy directed against the largest subclone*), over a wide range of scenarios encompassing literature and clinical experience across oncology^6^.

DPM in principle requires high resolution data on subclonal composition at multiple timepoints, potentially leading to a high-cost paradigm, as well as one that is invasive, given that sensitivity for rare subclones in liquid biopsies might be limited. Hence, we asked whether 2 moves of DPM provided patient benefit, and how this might compare to the benefit from 40 moves over a 5-year period.

Moreover, the average benefit of DPM was driven by 31% of the virtual patients who experienced significant benefit while the other 69% of patients had equivalent outcomes on CPM and DPM. Hence it is important to identify the subset of patients who will benefit from DPM.

This manuscript also concerns the development of an ***evolutionary classifier (EC)*** to select this important patient subset. In principle this could be accomplished by direct calculation using the DPM equations over an entire clinical course, but that approach may not be as robust in clinical situations in which the available data may be incomplete compared to ideal DPM input data, particularly if data are available only at a small number of early timepoints. The approach we have discovered for identifying patients who will benefit may be readily generalizable to other EGPM approaches that might ideally require detailed molecular data at multiple timepoints. This would greatly facilitate clinical testing of EGPM approaches.

In the results section, we will describe the development and performance of the evolutionary classifier, as well as surprising results for a 2-move DPM window vs. the original 40-move paradigm. In the discussion, we will address the impact and innovative aspects of these findings and their possible generality. We will also discuss limitations of the work and future plans.

## METHODS

### Clinical evaluation of DPM

Clinical testing of EGPM faces several hurdles that may affect the ability to get complete input datasets^6^. Highly sensitive and specific non-invasive assays to detect rare subclones and predict their drug sensitivity and resistance properties and evolutionary dynamics are ideally required. However, these concerns are out of the scope of this manuscript, which instead focuses on selecting patients assuming the relevant input parameters are (to some degree) available, and thus enhancing the efficiency of clinical studies to evaluate the merit of EGPM. The EC developed in this work may be more robust in settings where data are available at early timepoints only, compared to direct computation using the DPM equations.

Clinical studies of highly adaptive EGPM algorithms that consider multiple therapies face a challenge. Highly adaptive EGPM presents a very large decision tree of therapy sequences which will be assigned to patients individually based on their initial state and evolutionary dynamic parameters^11^, and will in real applications need periodic reassignment if future states diverge from predictions. In DPM, which takes into account possible need for dose reduction in simultaneous combinations, if there are two partially non-cross resistant therapies A and B, there are three options at each move: full dose A, full dose B, and a simultaneous combination of A and B at reduced dose. (We note a “therapy” may itself be a combination of drugs. A “therapy” is defined as a drug or group of drugs targeted against a particular subclone). Due to frequent adaptation, both A and B can be delivered at high intensity early on, even if administered as monotherapy pulses. The number of possible paths if two therapies are considered over n moves is thus 3^n^. If we consider only 2 moves, or a 12-week window, there are 9 sequences to consider.

### Evolutionary classifier (EC)

In contrast to a conventional biomarker classifier, which informs a matching between patients and optimal therapies, we seek to develop an evolutionary classifier that will match patients to strategic algorithms for computing optimal treatment sequences. Moreover, the classifier matches not just based on static molecular properties but based on dynamic properties such as growth and mutation rates that may predict future states. To support the translation of DPM into the clinic, we note the EC must now perform two classifications. EC1 must first identify patients most likely to benefit from DPM, who will then be the trial participants. EC2 must then sort patients selected by EC1 into optimal therapy sequences. Figure 1 shows how these two classifiers might function together in the context of a window pilot study in the neoadjuvant setting in ER/HER2 double positive breast cancer.

**Figure 1.**
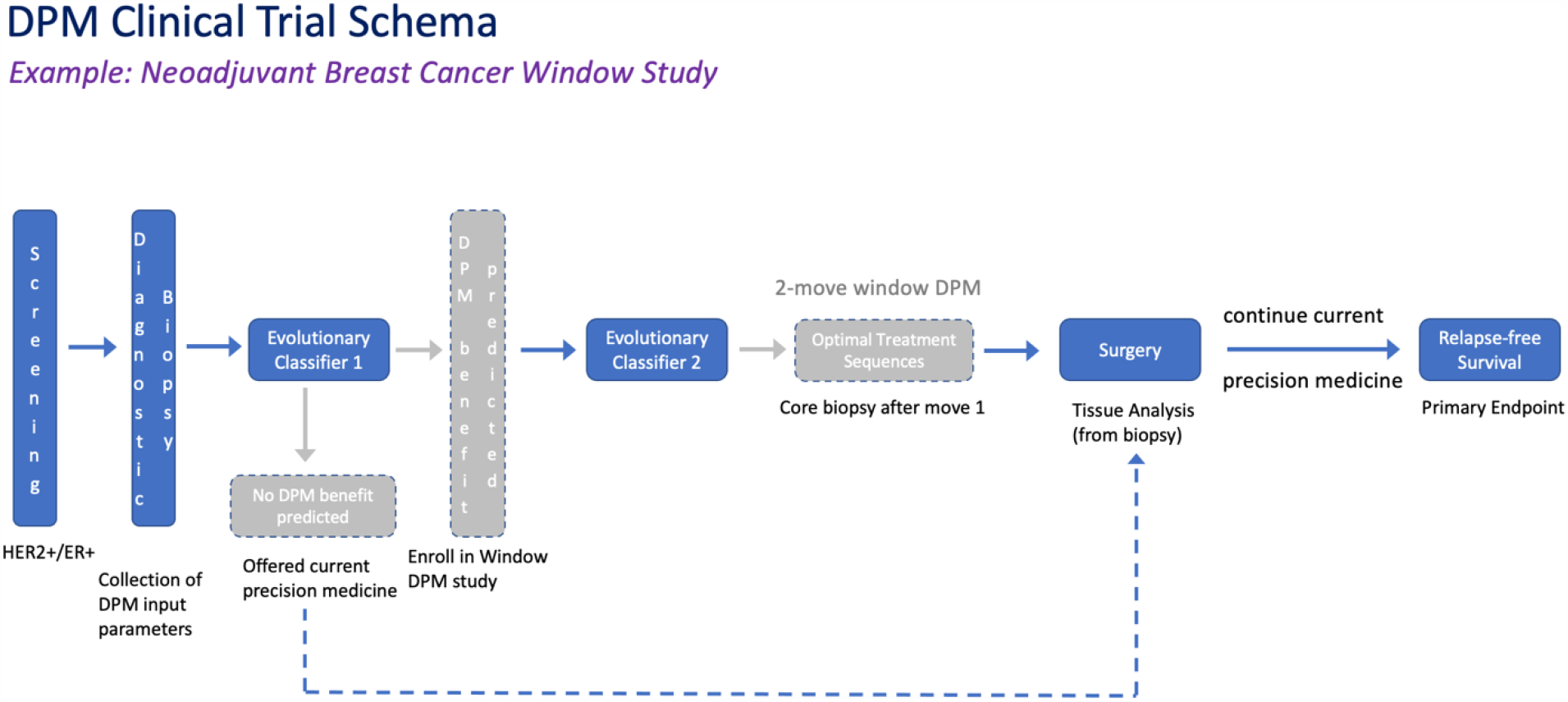
Vision of an evolutionary window pilot clinical trial using novel mathematical and computational tools of an evolutionary classifier (EC). Within the neoadjuvant setting, breast cancer patients are screened for double HER2 and ER positive status. A diagnostic biopsy collects the individual DPM input parameters required for EC1 predictions. Patients expected to benefit from DPM are enrolled in a 2-move window trial. EC2 assigns patients to the evolutionary treatment sequence over the 2 moves predicted to be optimal. Core biopsy is performed at the end of move 1. Another biopsy is performed at surgery for tissue analysis. Patients continue on CPM after surgery. Patients who are not predicted to benefit from DPM will simply receive CPM before and after surgery. The primary endpoint will be relapse-free survival in a high-risk group. Subsequent clinical development steps (not shown) would contain randomization between DPM and CPM.

DPM simulations were performed on nearly 3 million virtual patients comprising different combinations of input parameters as described in the introduction^6^. A system of ordinary differential equations based on the evolutionary model was integrated piecewise to simulate clinical time courses and DPM and CPM recommendations for each move read out to support EC1 and EC2 using strategy 2.2 to represent DPM and strategy 0 to represent CPM^6^. Strategy 2.2 prioritizes prevention of the birth of multiply resistant cells unless the total cancer burden exceeds a predetermined threshold, whereupon overall cytoreduction is prioritized. In practice, determining when it is critical to prioritize overall cytoreduction would rely on the judgement of the treating physician. Strategy 0 focuses on personalized cytoreduction by giving therapy directed at the most prevalent subclone. Death occurs in the simulation when cell numbers exceed a second predetermined threshold^6^.

Kaplan-Meier analyses and calculation of EC1 statistical properties were performed using functions in R.

## RESULTS

### Nearly equal benefit from Full DPM and 2-Move Trial DPM

In order to consider a window study design in which DPM is given only within a 2-move window, we must determine the benefit conferred by only 2 moves of DPM. Figure 2A shows the surprising finding that the first 2 moves of DPM confer most of the benefit of full DPM. In the entire simulation, 40 moves of DPM confers a hazard ratio, HR (DPM/CPM), of 0.52 and 2 moves of DPM confers a HR of 0.55.

**Figure 2.**
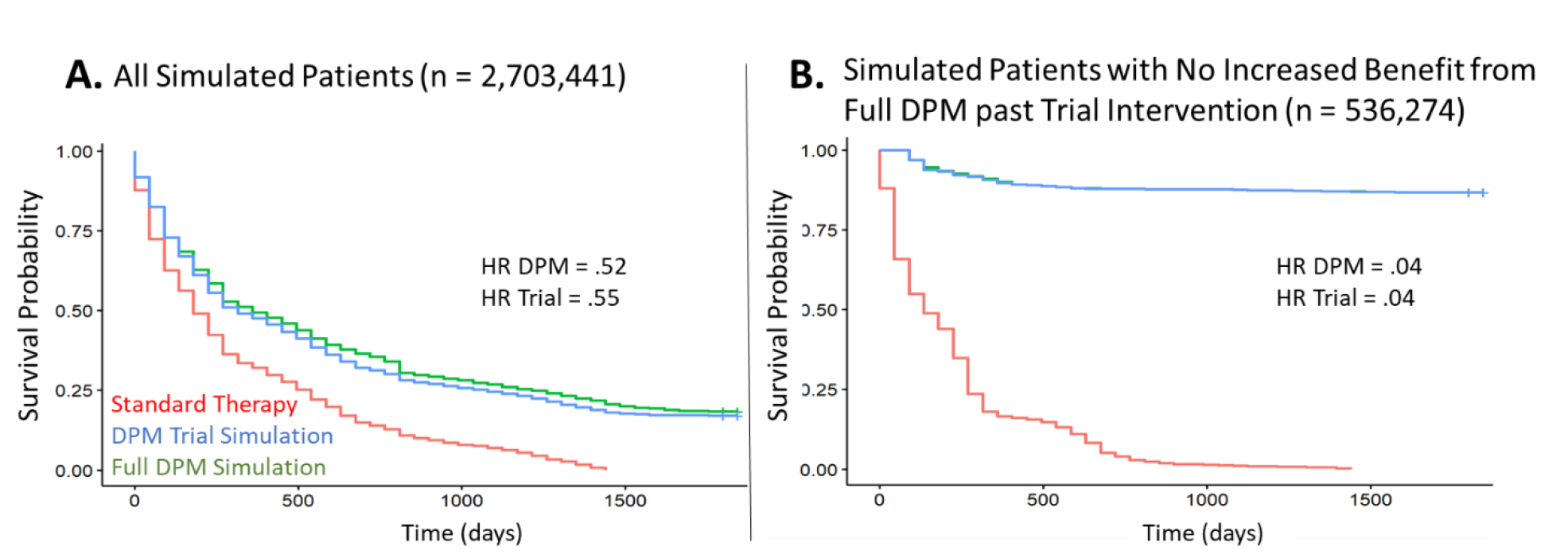
(A) Kaplan-Meier plot of simulated survival, which is increased over the standard therapeutic sequence (red) when following the strategy defined by DPM for the entire clinical course (full DPM, green) or the first two moves only (DPM trial simulation, blue). Benefits from trial DPM and full DPM are similar, as shown by the hazard ratios. (B) Kaplan-Meier plot of those virtual patients predicted to have similar survival (within 25% relative and 60 days absolute) between trial DPM and full DPM. This group of patients, about 20% of those from the original population, experience substantial benefit from either trial DPM or full DPM. (Note p values in figures A and B (not shown) are highly significant, but with such large sample sizes p values would be significant even with small differences. Similarly, 95% confidence intervals would be extremely tight due to the large number of virtual patients, not truly reflecting the total uncertainty inherent in such an analysis).

### Extraordinary benefit for virtual patients whose benefit from full DPM and 2-move trial DPM are closely equal

Figure 2B shows the benefit predicted for those virtual patients whose survival on trial DPM and full DPM differs by less than 25% relative and less than 2 months absolute. These patients, representing nearly 20% of the original population, experience an HR of 0.04.

### Construction of EC1 and its performance

Given that the first two moves of DPM appear to confer most of its benefit, we hypothesized that DPM would not be beneficial compared to CPM in patients with specific parameter configurations where the recommendations for both moves 1 and 2 were identical between DPM and CPM.. Thus, a patient was labelled by EC1 as potentially benefitting from DPM if DPM and CPM diverged in one or both of the first two moves. We can evaluate the performance of EC1 by comparing the results of CPM and DPM, where a patient was defined as benefitting from DPM in simulation if it provided at least 25% relative and 2-month absolute survival advantage compared to CPM. The performance of EC1 as a predictor of benefit is shown in Table 1, where the high sensitivity and negative predictive power demonstrate its effectiveness at identifying at identifying a significant portion of the simulated population that should not be enrolled in the 2-move trial of DPM. Figure 3 shows Kaplan-Meier curves of three treatment strategies – CPM, full DPM (applying the DPM algorithm over the full 40-move course), and trial DPM (applying DPM to the first two moves and reverting to CPM afterward) – among the patients with identical recommended moves 1-2 between DPM and CPM. No clinically significant advantage of DPM is seen under these conditions. Note p values are not shown in this research brief since the sample size in the virtual trial is extremely large, conferring statistically significant p values even in the face of clinically insignificant differences.

**Table 1**. Statistical performance of an initial heuristic EC1 based on differences between DPM’s and CPM’s recommended sequence of two drugs or a combination for the first two moves. Benefit is defined by an overall increase in survival by at least 25%, and at least 60 days when using the DPM approach.

**Figure 3.**
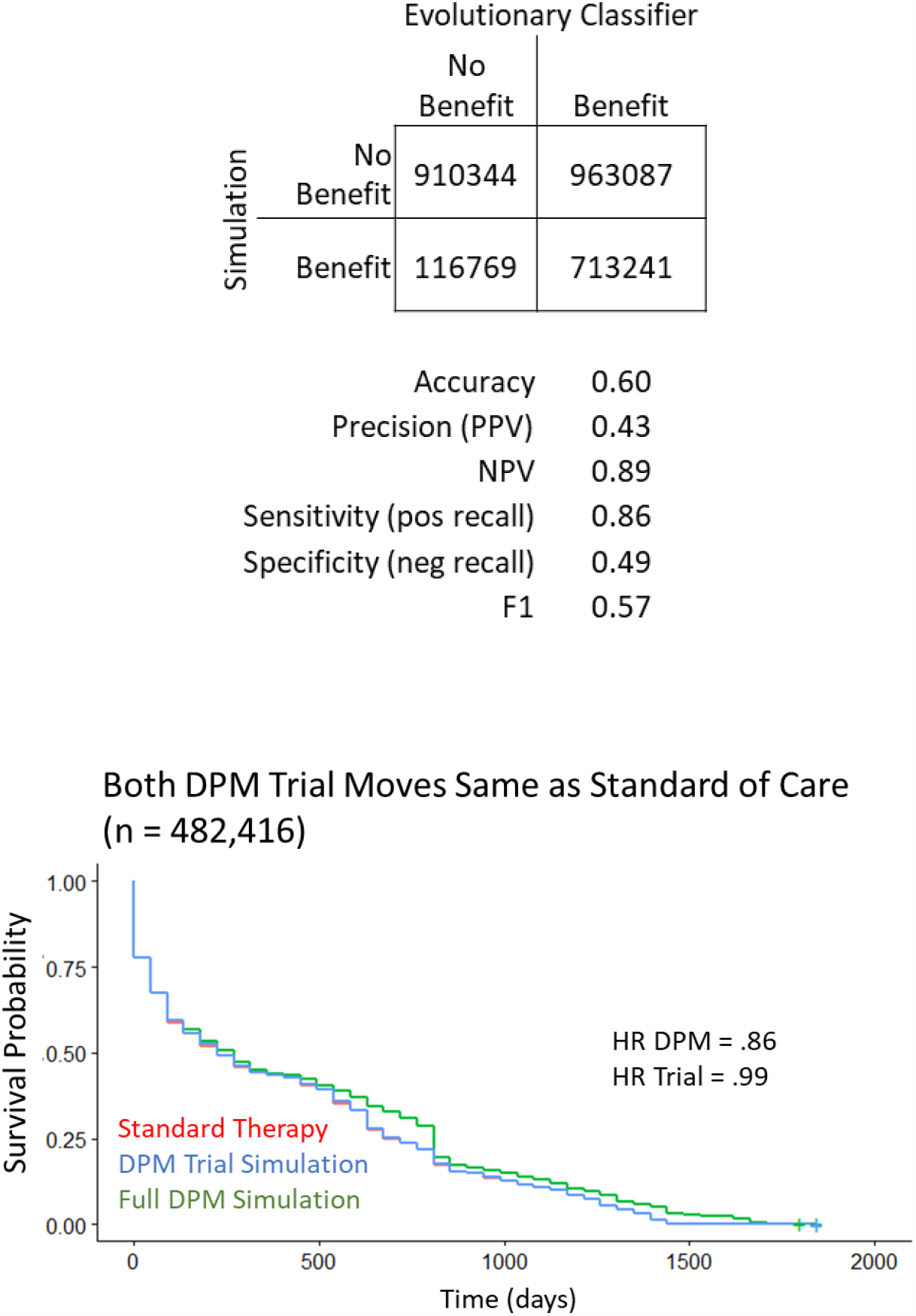
Kaplan-Meier Plot for a simulated trial of DPM, where three treatment strategies were compared. CPM (Standard Therapy) (red) recommendations are based on the therapy that provides the greatest response to the largest population of tumor cells, and the Full DPM (green) recommendations minimize the emergence of multiple drug resistance at the expense of overall tumor shrinkage. The DPM Trial (blue) treatment strategy used the DPM recommendations for the first two therapeutic interventions before reverting to the standard therapeutic strategy. The resulting populations can be grouped based on comparing the recommendations of DPM and CPM. Here we show that if DPM and CPM give identical recommendations for moves 1 and 2, there is little to no benefit of DPM. If only move 2 differs, DPM gives a clinically significant benefit (hazard ratio 0.74), but trial DPM does not (hazard ratio 0.86) (data not shown). (Note 95% confidence intervals (not shown) are tight due to the large number of virtual patients, but this may not truly reflect the total uncertainty inherent in such an analysis).

## DISCUSSION

Previously, biomarker classifiers have been used to categorize patients and match them to therapy according to conventional precision medicine. These classifiers have consisted of a single biomarker such as a mutation, or a panel of biomarkers such as a gene expression profile, involving categorical or continuous variables, the latter converted to categorical by cutoffs^12-16^. Here, the classifier is being applied to determine if patients will benefit from DPM and to match patients to individualized therapy sequences of the same drugs according to an algorithm based on initial subclonal state variables and evolutionary dynamics parameters^6^. The EC input values are thus unique, related in a complex way by a system of differential equations^6^, and deployed for unique purposes compared to previous biomarker classifiers.

The classification, however, can be dramatically simplified by a high-level principle. Populations can be enriched for DPM benefit based on comparison of therapy recommendations by DPM and SOC in just the first move. It is striking that this is true for DPM, which has the ability to plan each move against a future horizon far longer than the proposed window, i.e. up to 5 years^11^.

DPM and potentially other EGPM approaches require repeated observations over time at deep sub-clonal resolution. While single cell methods are improving, they are expensive and sensitivity and accuracy may be insufficient to distinguish rare cells from technical error. According to DPM simulations, one cell in 100,000 can alter the optimal strategy if it is an extreme hypermutator^6^. These challenges are magnified for liquid biopsies which are nonetheless essential to minimize patient risk and inconvenience if prolonged monitoring is required. The finding that a brief window of DPM provides similar benefits to prolonged DPM opens up the possibility of reduced patient risk and inconvenience, and reduced cost. The proposed 3-month window may allow DPM to be deployed in high risk neoadjuvant settings, where tissue availability at diagnosis and at surgery are routine and only one additional core biopsy between these timepoints would be required.

From the point of view of dynamic models of cancer treatment, especially EGPM approaches, it also raises the question of how general this high-level approach to classifying patients can be. Can patients who will benefit from other treatment approaches based on other dynamic models of cancer be selected based on whether the recommended treatment differs from SOC in an initial treatment window, and what in a given model determines the required length of this window?

One might imagine, in patients who benefit equally from a finite number of moves directed by EGPM and more extended EGPM, that the initial moves convert the patient’s cancer to a state similar to the initial state of a patient who cannot benefit. Once these states precluding further benefit are identified, periodic checks can in principle be instituted to govern stopping and restarting of an EGPM as needed, again optimizing cost-effectiveness. While details will differ for each EGPM, the high-level principle may be general.

Future work is necessary to more deeply understand these findings. We will determine whether this process can in fact be used to stop and restart DPM. We will create subsets of the very large simulation population based on recommended moves and on clustering based on initial state variables and input parameters, and visualize and analyze representatives of each cluster to elucidate underlying evolutionary mechanisms for DPM benefit. There may be multiple mechanisms behind significant DPM benefit in the nearly one million virtual patients who demonstrated it.

## Data Availability

The data from the study is derived from computational simulations, the methods for their generation and recreation are provided in Beckman et. al., PNAS (2012) https://doi.org/10.1073/pnas.1203559109

## ACKNOWLEDGEMENT

We thank Dr. Jin Young Yang for assistance with DPM simulations. We thank Dr. Rebecca Riggins for manuscript review, and the Academia Sinica Data Science Statistical Cooperation Center for assistance. This work is supported by The Royal Society International Exchanges Award IES\R3\183092 to DP and RAB and the US Department of Defense Breast Cancer Research Program Breakthrough Award W81XWH-20-1-0760 to RAB.

